# Clarifying Cognitive Control Deficits in Psychosis via Drift Diffusion Modeling

**DOI:** 10.1101/2023.08.14.23293891

**Authors:** Chen Shen, Olivia L. Calvin, Eric Rawls, A. David Redish, Scott R. Sponheim

## Abstract

Cognitive control deficits are consistently identified in individuals with schizophrenia and other psychotic psychopathologies. In this analysis, we delineated proactive and reactive control deficits in psychotic psychopathology via hierarchical Drift Diffusion Modeling (hDDM). People with psychosis (PwP; N=123), their first-degree relatives (N=79), and controls (N=51) completed the Dot Pattern Expectancy task, which allows differentiation between proactive and reactive control. PwP demonstrated slower drift rates on proactive control trials suggesting less efficient use of cue information for proactive control. They also showed longer non-decision times than controls on infrequent stimuli sequences suggesting slower perceptual processing. An explainable machine learning analysis indicated that the hDDM parameters were able to differentiate between the groups better than conventional measures. Through DDM, we found that cognitive control deficits in psychosis are characterized by slower motor/perceptual time and slower evidence-integration primarily in proactive control.

## Introduction

Cognitive control is the ability to regulate, coordinate, and sequence thoughts and actions in accordance with internally maintained behavioral goals (Braver, 2012). Deficits in cognitive control are central to schizophrenia and limit the daily functioning of people with this disorder (Alptekin et al., 2005; Pascal de Raykeer et al., 2019; Ueoka et al., 2011). Among all cognitive functioning deficits, deficits in cognitive control have the strongest effect on quality of life (Savilla et al., 2008), which makes it an essential target for intervention. Deficits of cognitive control in psychotic psychopathology are often characterized by difficulties in incorporating information about the current context to determine a correct response (i.e., proactive control; Braver et al., 2001, 2009). Experimental tasks used to measure cognitive control deficits vary widely but share the requirement of responding adaptively to stimuli that are incompatible with prepotent or automatic response tendencies. For example, the Stroop test (Stroop, 1935) and the Flanker test (Eriksen & Eriksen, 1974) are characterized by their within-stimulus conflicts. The AX-Continuous Performance Test (Cohen et al., 1999) and its non-letter variant the Dot Pattern Expectancy Task (DPX; MacDonald et al., 2005) were also developed to capture cognitive control. These tasks temporally separate the within-stimulus conflicts into “cue” (which defines a context) and “probe” periods (Figure 1A). The stimulus sequences allow the participant to engage in proactive control by anticipating the correct response from the cue and/or reactive control by determining the correct response from the probe.

**Figure 1.**
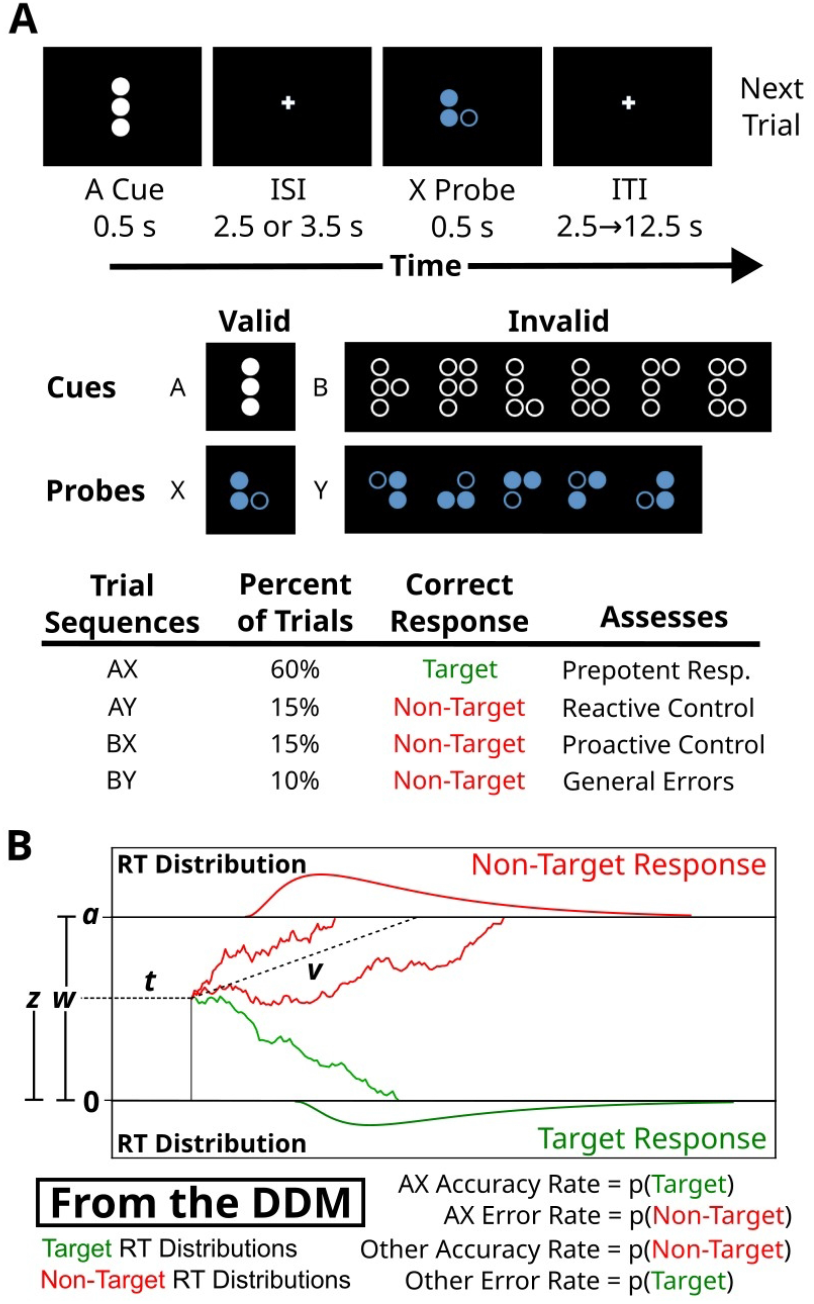
**A**. The Dot Pattern Expectancy (DPX) Task. Each trial consists of a “cue” stimulus presentation followed by a “probe” stimulus after a brief interstimulus interval (ISI). The stimuli consist of dot patterns with there only being one valid cue (A) and one valid probe (X) amongst similar invalid stimuli (B and Y). The most frequent cue-probe sequence is AX, which promotes a response prepotency on A cues for the target response. After each trial there is a variable intertrial interval (ITI). **B**. The drift diffusion model (DDM). Four parameters are used by drift diffusion models to fit the reaction time and error performance of the subject. These parameters are the bias (*z*), the decision threshold (*a*), the drift rate (*v*), and the non-decision time (*t*). These four parameters produce reaction time and response probabilities for the subjects’ target and non-target responses. The ratio of the bias and decision threshold (*z*/*a*) is typically fit via hierarchical DDM, which is denoted here as *w*.

Behavior on the AX-CPT and DPX tasks has been most frequently characterized using the participant’s accuracy, average reaction times on correct trials, and indices based on these features. The most commonly used index is d’-context, which assesses context processing via accuracy rates (Servan-Schreiber, 1996). A separate metric that was developed to assess proactive cognitive control is the proactive behavioral index (PBI), which can be derived from either reaction time or accuracy data (Braver et al., 2009). Deficits in cognitive control among people with schizophrenia have been indicated by greater BX error rates (Poppe et al., 2016; Stephenson et al., 2020), prolonged average reaction times (Lesh et al., 2013; Stephenson et al., 2020), and lower d’-context scores (Jones et al., 2010; Poppe et al., 2015). Relatives of people with schizophrenia often demonstrate attenuated deficits in these measures (Delawalla et al., 2008; MacDonald et al., 2003; Poppe et al., 2015; Reilly et al., 2016; Richard et al., 2013). While these approaches are adequate for describing trends in the central tendencies of reaction times and accuracy, it omits within-subject variation in responding which contains valuable information about underlying cognitive processes. Overall, these analyses treat accuracy and reaction times as independent components that are best assessed via central tendencies, but these indices are actually intertwined behavioral outcomes with informative within-subject variation.

Accuracy and reaction times can be addressed in a singular analysis that encompasses within-subject variability via drift-diffusion modeling (DDM; Figure 1B). In DDMs, an underlying cognitive process noisily integrates evidence to produce an observable response. The DDM characterizes the decision-making process with 4 parameters that represent the rate at which evidence is accumulated (drift rate, *v*), how much evidence is required to act (decision threshold, *a*), the degree of prestimulus bias towards a particular response (bias, *w*), and the response delay due to non-decision related processes (non-decision time, *t*). Given that both longer reaction times and higher error rates are observed among people with psychosis in the AX-CPT and DPX tasks (Jones et al., 2010; Lopez-Garcia et al., 2013; MacDonald & Carter, 2003; Smucny et al., 2019; Stephenson et al., 2020), we suspected that people with psychosis could markedly differ from control subjects in how they integrate evidence to produce a response.

Previous DDM analyses of the behavior of people with schizophrenia have most often observed deficits in drift rates and non-decision times, but not always (Gupta et al., 2022). Specifically, people with schizophrenia showed longer non-decision times and slower drift rates on the N-back (Fish et al., 2018) and digit-symbol-like coding tasks (Mathias et al., 2015). On a reward-punishment task, Moustafa et al. (2015) observed greater non-decision times, but with greater decision thresholds instead of the typically observed difference in drift rate. Some recent analyses by Smucny and colleagues have suggested slower drift rate in individuals with recent-onset schizophrenia during reward anticipation (Smucny, Hanks, Lesh, O’Reilly, et al., 2023), and cognitive control on the AX-CPT (Smucny, Hanks, Lesh, & Carter, 2023). It is important to note that Smucny and colleagues’ analytic approach is somewhat limited and cannot resolve differences in non-decision times or bias that are likely to exist. The only study that showed a difference in biases was on a temporal prediction task (Limongi et al., 2018). In the only analysis of relatives of people with schizophrenia via DDM, Fish et al. (2018) identified slower drift rates and longer non-decision time in siblings of individuals with schizophrenia than controls on a sustained attention task. Recent simulations of the effects of excitation-inhibition imbalance also suggest utility in applying DDM to cognitive control tasks in people with psychosis (Calvin & Redish, 2021; Lam et al., 2022). From the perspective of these simulations, the longer non-decision times and slower integration rates observed in schizophrenia could be attributed to deviations in the efficacy of glutamate (excitation) and GABA (inhibition) that cause weak neural representations of stimuli within circuits that are necessary for appropriate responding. This mix of findings and the potential effects of excitation-inhibition balance on reaction times suggested that analyzing cognitive control via DDM would provide insights into psychosis.

We applied hierarchical DDMs (hDDM) to DPX reaction time and response data obtained from PwP, their first-degree biological relatives, and controls. Our goal was to directly examine how aspects of the cognitive control processes differ across these groups during proactive and reactive control. We hypothesized that people with psychosis would have longer non-decision times and slower drift rates, and that this would be attenuated in relatives. We also hypothesized that there may also be a group difference in the bias parameter given the identified deficits in proactive control (i.e., context processing) in individuals with psychotic psychopathology.

## Results

### Participants

We reanalyzed DPX reaction time data that was collected during the Psychosis Human Connectome Project (see Demro et al., 2021 for details). In summary, the study recruited 253 participants, which we separated into three groups. The groups were 123 people with psychosis (PwP), 79 of their first-degree biological relatives, and 51 controls. The groups differed in their ages and sex distributions (Table 1) so we controlled for age, sex, and, in addition, familial relationships in analyses. For additional details please see the Methods section.

**Table 1.** Demographics and Clinical Information of the Participants. PwP=People with a history of psychosis. *Fisher’s exact test. Parent education: median of max of parents’ education; (1: 7th grade or less; 2: between 7th and 9th grade; 3: between 10th and 12th grade; 4: high school graduate/GED; 5: partial college; 6: college graduate; 7: graduate degree) **Kruskal-Wallis rank sum test. Significant post-hoc group differences: a=relatives vs. both controls and PwP; b=controls vs. relatives vs. PwP, c=PwP vs. both controls and relatives.

|  | Controls<br>(N=51) | Relatives<br>(N=79) | PwP<br>(N=123) | Tests |
| --- | --- | --- | --- | --- |
| **Diagnoses/**<br>**Diagnosis of**<br>**Relative (N)** | N/A | Bipolar w/<br>psychosis (23)<br>Schizoaffective<br>disorder (9)<br>Schizophrenia<br>(47) | Bipolar w/<br>psychosis (35)<br>Schizoaffective<br>disorder (15)<br>Schizophrenia<br>(73) | N/A |
| **Age (SD)** | 38.4 (13.1) | 44.8 (14.8) | 37.3 (12.2) | $F_{(2,250)}=8.26, p<.001^a$ |
| **Female N (%)** | 25 (49%) | 52 (66%) | 52 (42%) | $\chi^2_{(2)}=10.77, p=.005$ |
| **Race (N)** | | | | $*p=.29$ |
| *American Indian or*<br>*Alaskan Native* | 0 | 0 | 1 |  |
| *Asian or Pacific*<br>*Islander* | 1 | 1 | 6 |  |
| *Black, not of*<br>*Hispanic Origin* | 3 | 5 | 20 |  |
| *Hispanic* | 1 | 2 | 4 |  |
| *White, not of*<br>*Hispanic Origin* | 45 | 69 | 88 |  |
| *Other* | 1 | 2 | 4 |  |
| **Edu (Years)** | 16.2 (2.5) | 15.2 (2.2) | 14.1 (2.1) | $F_{(2,249)}=16.96, p<.001^b$ |
| **Parent Education** | 6 | 6 | 5 | $**\chi^2_{(2)}=4.94, p=.08$ |
| **Estimated IQ (SD)** | 106.7 (11.0) | 102.3 (10.8) | 98.2 (11.4) | $F_{(2,250)}=10.99, p<.001^c$ |
| **BPRS Total (SD)** | 27.8 (4.2) | 32.5 (6.7) | 45.2 (12.6) | $F_{(2,250)}=74.19, p<.001^b$ |
| **SPQ Total (SD)** | 7.7 (7.9) | 14.4 (12.7) | 29.9 (15.3) | $F_{(2,250)}=62.68, p<.001^b$ |

### Conventional measures of DPX

#### Error rates

Groups differed in their error rates (Figure 2A). A linear mixed effects regression (LMER) model of error rates revealed a main effect of group (*F*_(2,245.93)_=3.53, *p*=.03), a main effect of trial stimulus sequences (*F*_(3,747.10)_=24.91, *p*<.001), and an interaction between group and trial sequence (*F*_(6,747.12)_=2.38, *p*=.03). Consistent with past literature, PwP made more errors on BX trials than controls (*t*_(680)_=3.57, *p*=.001) and relatives (*t*_(632)_=-3.36, *p*=.002), while controls and relatives did not differ on BX error rates (*t*_(609)_=0.51, *p*=.87). The groups did not differ significantly on the other trial sequences (|*t*|s<1.57, *p*s>.26).

**Figure 2.**
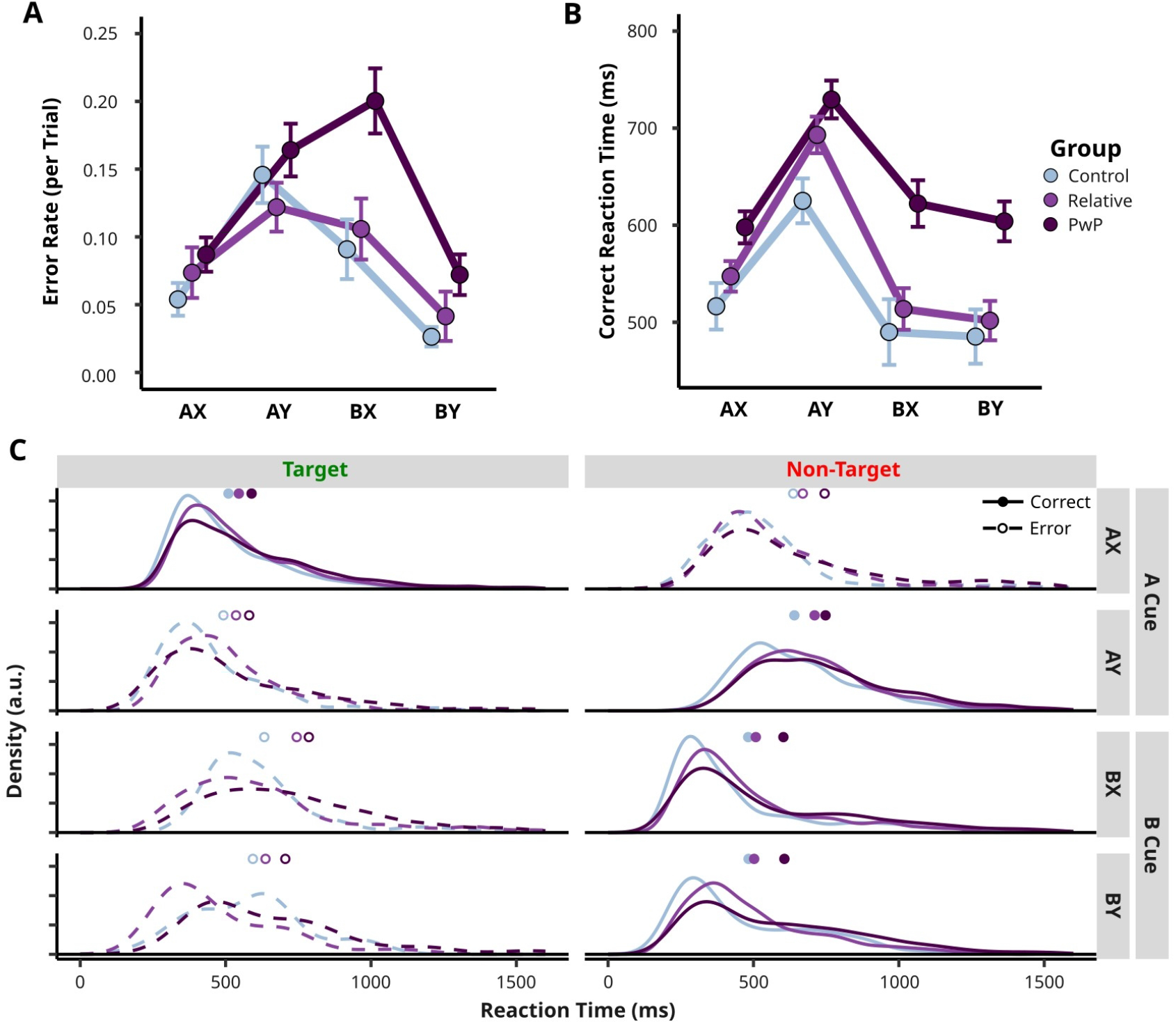
**A**. Error rate means and standard errors of participants in each group. **B**. Reaction time means and standard errors of participants in each group. **C**. The normalized reaction time probability density distributions for each group’s target and non-target responses. The dots above each of the panels indicate the participant mean reaction times from panel B.

#### Reaction times (RT)

Groups also differed in their reaction times (Figure 2B). A LMER model of the by-individual mean correct response reaction times showed a main effect of group (*F*_(2,155.75)_=9.24, *p*<.001), a main effect of trial sequence (*F*_(3,734.65)_=169.88, *p*<.001), and an interaction between group and trial sequence (*F*_(6,734.75)_=3.70, *p*=.001). PwP were slower on BX and BY trials compared to controls (*t*_*BX*(337)_=-3.80, *p*<.001, *t*_*BY*(336)_=-3.70, *p*<.001) and relatives (*t*_*BX*(269)_=3.89, *p*<.001, *t*_*BY*(269)_=4.04, *p*<.001). PwP were significantly slower on AY trials (*t*_(335)_=-3.36, *p*=.003) and AX trials (*t*_(335)_=-2.38, *p*=.047) than controls.

#### d’-context and Proactive Behavioral Index (PBI)

One-way ANOVAs revealed group differences in d’-context scores (*F*_(2,226.28)_=7.36, *p*<.001) and PBI-RT (*F*_(2,178.05)_=4.59, *p*=.01), but not in PBI-accuracy (*F*_(2,173.79)_=1.62, *p*=.20). PwP demonstrated lower d’-context scores and PBI-RT than both relatives (*t*_*d’-context*(235)_=-3.15, *p*_*d’-context*_=.005; *t*_*PBI-RT(*181)_=-2.41, *p*_*PBI-RT*_=.04) and controls (*t*_*d’-context*(248)_=3.07, *p*_*d’-context*_=.007; *t*_*PBI-RT*(236)_=2.40, *p*_*PBI-RT*_=.04), suggesting less efficient context processing and less utilization of proactive control. The control and relative groups did not differ from each other on d’-context scores or PBI-RT (*t*_*d’-context*(227)_=0.175, *p*_*d’-context*_=.98; *t*_*PBI-RT*(238)_=0.28 *p*_*PBI-RT*_=.96).

### DDM Parameters

The group reaction time distributions indicated that the participant’s reaction times were poorly described by their mean reaction times, and suggested that there were significant group differences in the underlying evidence integration process (Figure 2C). Thus, we hierarchically fit DDMs to the participant behavior to get individual estimates of the drift-rate (*v*), decision threshold (*a*), non-decision time (*t*), and the response bias (*w*) (Figure 1B). We found that the maximally flexible model (Figure 3A), which allowed *a, t*, and *v* parameters to vary by trial sequence and *w* by the cue type, was the best description of participant reaction times and responses. All of the best models showed good convergence via Gelman-Rubin with ^*R*s < 1.2 on all individual parameters.

**Figure 3.**
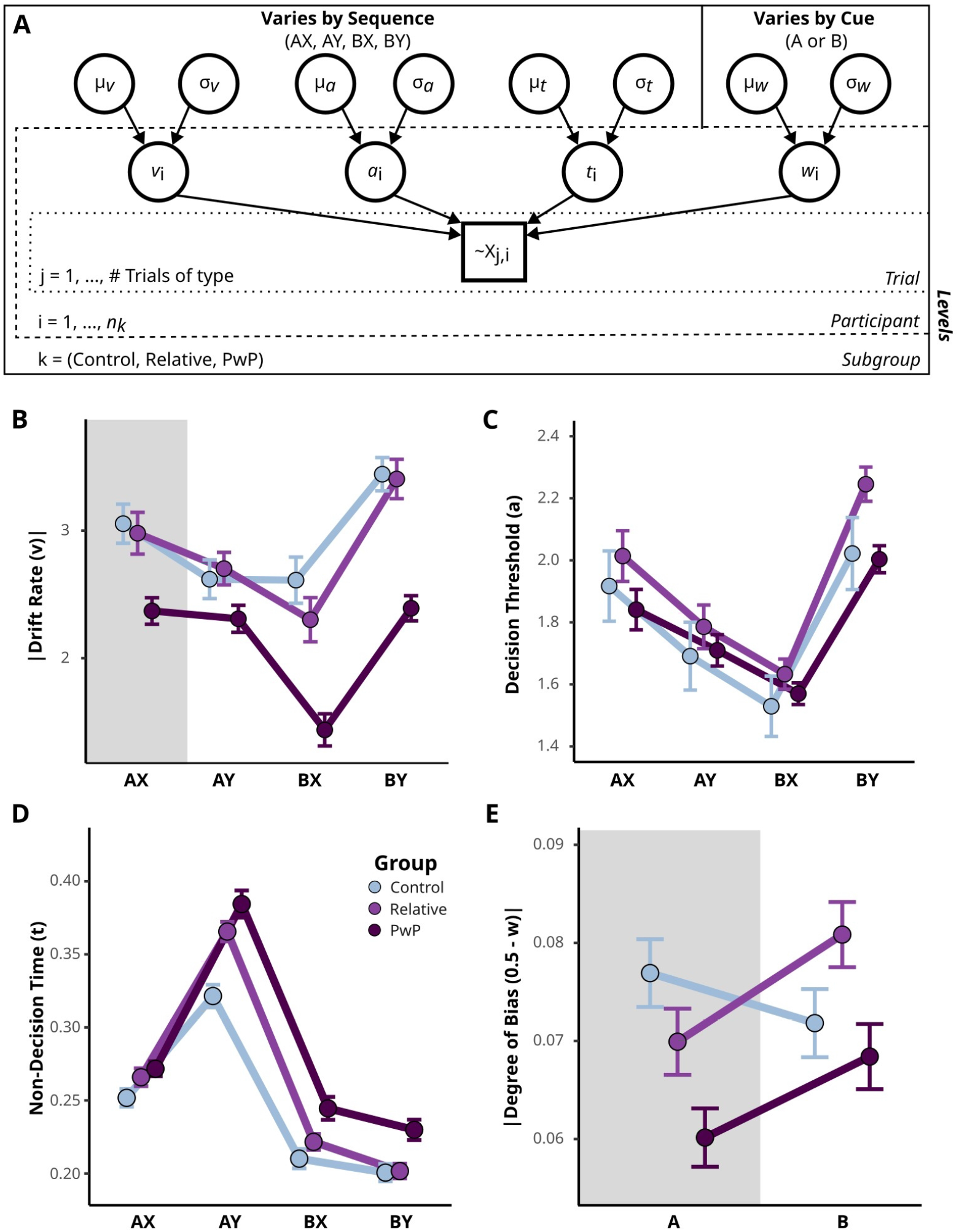
**A**. Diagram of the best fitting hierarchical drift diffusion model (hDDM) to the distribution of participant responses across trials (∼**X**_**i,j**_). The best fitting hDDM permitted the rate-of-integration, decision threshold, and non-decision time to vary by trial type (i.e., 1 of each parameter for AX, AY, BX, and BY) and the bias to vary by the cue (i.e., 1 parameter for A cues and 1 for B cues). μ and σ are the subgroup means and standard deviations of the parameters that are denoted in their subscripts. Each subgroup was independently fitted. **B**. The means and standard errors of the drift rate parameter by group. The gray section indicates that the sign of the parameter was negative (i.e., towards a target response) prior to taking its absolute value. **C**. The means and standard errors of the decision threshold by group. **D**. The means and standard errors of the non-decision time threshold by group. **E**. The means and standard errors of the degree of bias. The gray section indicates that the sign of the calculated parameter was negative (i.e., towards a target response) prior to taking its absolute value. The absolute value was taken after calculating the mean and standard errors.

#### Drift Rate (*v*)

For the sake of comparing across all trial stimulus sequences, we made the direction of AX trial drift rates positive (the only target trial) (Figure 3B). A LMER model examining drift rate across group and trial sequences revealed main effects of group (*F*_(2,237.73)_=19.42, *p*<.001) and trial sequence (*F*_(3,749.02)_=38.22, *p*<.001), as well as an interaction between group and trial sequence (*F*_(6,749.04)_=3.71, *p*=.001; Figure 3B). PwP showed lower rates of information integration than controls (AX: *t*_(695)_=3.13, *p*=.005; BX: *t*_(695)_=5.55, *p*<.001; BY: *t*_(696)_=5.03, *p*<.001) and relatives (AX: *t*_(646)_=-3.03, *p*=.007; BX: *t*_(646)_=-4.48, *p*<.001; BY: *t*_(647)_=-5.36, *p*<.001) across all trials, except for AY trials where PwP showed comparable drift rates to relatives (*t*_(646)_=-2.00, *p*=.11) and controls (*t*_(695)_=1.45, *p*=.32). This pattern of findings indicates a specific deficit in evidence integration when PwP can utilize proactive control, but not when they are utilizing reactive control. This difference is particularly exacerbated when the cue stimulus provides *definitive* information about the correct response (i.e., B cue trials), rather than *suggestive* information (i.e., A cue trials). Relatives did not significantly differ from controls on drift rates across all trials (|*t*|s<1.41, *p*s>.34).

#### Decision Threshold (*a*)

The groups did not significantly differ in their decision thresholds (Figure 3C). An LMER model of decision threshold revealed a main effect of trial sequence (*F*_(3,749.03)_=69.85, *p*<.001), but no main effect of group (*F*_(2,156.80)_=0.43, *p*=.65) or interaction between group and trial sequences (*F*_(6,749.04)_=1.35, *p*=.23). Each pairwise comparison among the trial sequences was statistically significant (|*t*|s>3.99, *p*s<.001). Generally, the incongruent trial sequences (i.e., AY and BX sequences, which have mixed valid and invalid stimuli) had the lowest decision thresholds.

#### Non-Decision Time (*t*)

There were interesting group differences in the non-decision time (Figure 3D). An LMER model revealed a main effect of trial sequence (*F*_(3,749.10)_=402.68, *p*<.001), a main effect of group (*F*_(2,211.94)_=12.47, *p*<.001), and an interaction between group and trial sequence (*F*_(6,749.11)_=3.53, *p*=.002). The groups did not differ significantly on AX trials in their non-decision times (|*t*| s<1.94, *p*s>.13). However, PwP showed longer non-decision times than controls on AY (*t*_(634)_=-5.85, *p*<.001), BX (*t*_(634)_=-3.02, *p*=.007), and BY (*t*_(635)_=-2.68, *p*=.02) trials. This effect was most pronounced on AY trials, which require reactive control. The longer non-decision times on trials with a B cue suggests that there may be less proactive response planning when there is definitive information on the correct response compared to control participants. Relatives showed longer non-decision times than control participants on AY trials (*t*_(566)_=-2.99, *p=*.008). Relatives also showed shorter non-decision time on AY (*t*_(588)_=2.86, *p*=.01), BX (*t*_(588)_=3.05, *p*=.007) and BY (*t*_(589)_=3.74, *p*<.001) trials than PwP.

#### Degree of Bias (*w*)

To better equate differences in bias we adjusted the value such that it was an estimate of the deviation from indifference (i.e., 0.5) (Figure 3E). An LMER model on degree of bias revealed a main effect of group (*F*_(2,206.59)_=3.09, *p*=.047), no main effect of the cue (*F*_(1,250)_=1.70, *p*=.19), and no interaction between group and trial sequence (*F*_(2,250)_=1.43, *p*=.24). Although there was a main effect of group, post-hoc pairwise comparisons did not survive multiple comparison correction. PwP showed a lower degree of bias than their relatives (*t*_(218)_=-2.18, *p*=.076), but this did not reach significance since the test was two-tailed. PwP did not differ from controls on degree of bias (*t*_(245)_=1.78, *p*=.17).

### Classification Models to Determine Utility

We applied eXtreme Gradient Boosting (XGBoost) (Chen & Guestrin, 2016) model classification to the 14 DDM parameters, followed by a Shapley interpretation (SHAP). This analysis helps identify the presence of heterogeneity within a group in their evidence integration processes, and indicates which aspects of the evidence integration process are most uniquely important for differentiating between groups.

#### Controls vs. PwP

An XGBoost model classifying PwP from control participants had high discriminative performance, indicated by a cross-validated area-under-the-curve (AUC) of 0.91. The SHAP explanation of this model (Figure 4A) found that the most important variable in this model was B-cue bias, where increased bias, surprisingly, predicted a higher likelihood of being classified as a member of the PwP group. Other variables that were especially important to categorizing participants were v-BY, where lower drift rate predicted higher likelihood of being labeled PwP, t-AY, where longer AY non-decision times predicted greater likelihood of being labeled PwP, a-BX, where increased BX decision thresholds predicted greater likelihood of being labeled PwP, and t-BX, where longer non-decision times for BX trials predicted greater likelihoods of being labeled PwP.

**Figure 4.**
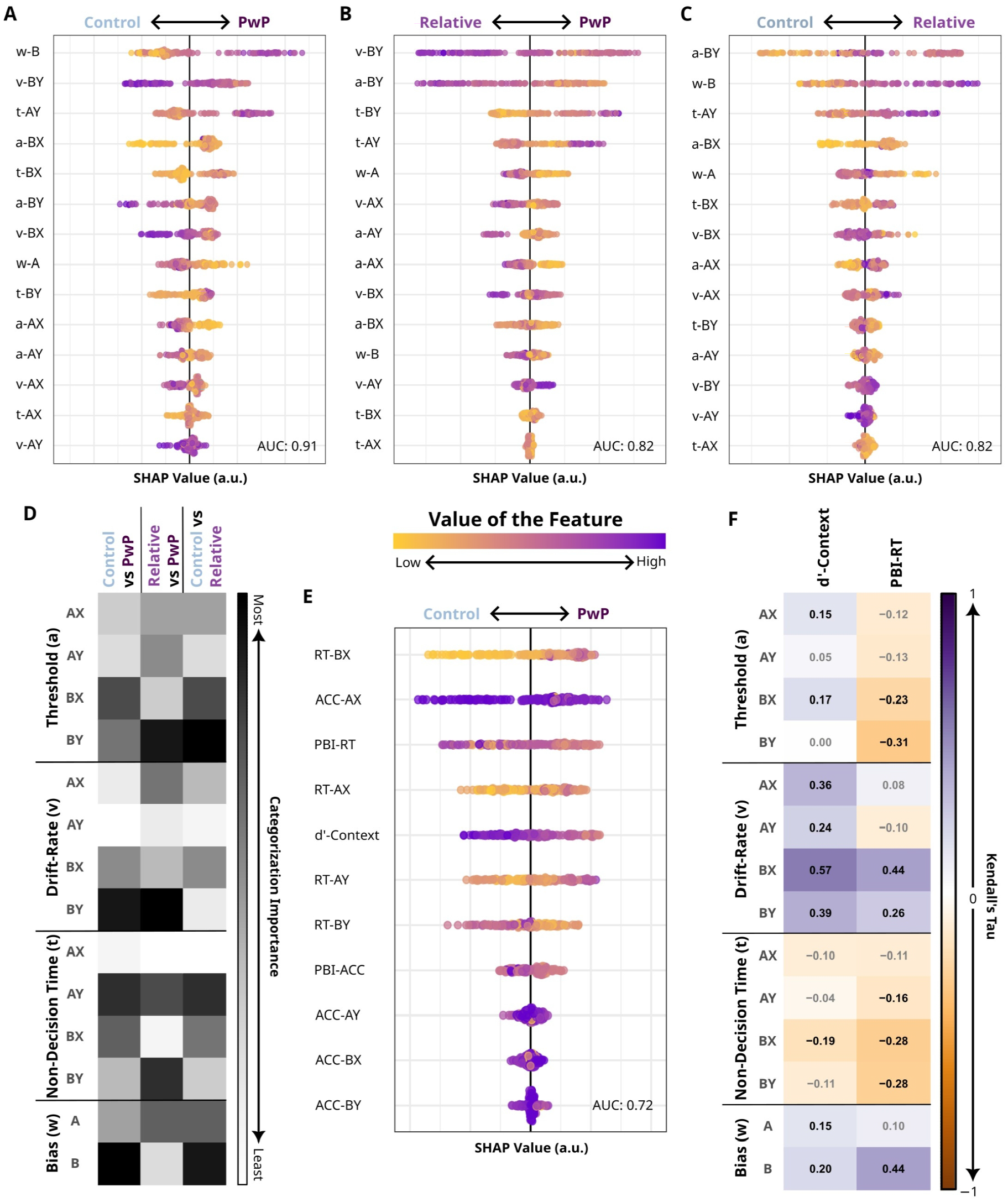
**A-C**. SHAP explanations of XGBoost model classifications. Each dot represents a single participant. Dots are arranged along the X-axis according to the impact each variable had on the model classification for each individual with the black line in the center indicating zero value. Dots to the left of the zero line indicate that the variable predicted membership in the class label to the left, and dots to the right similarly indicate that the variable predicted membership in the class label to the right. Parameters are listed in the order of classification importance. **D**. Summary of how important each parameter was for differentiation between the groups. **E**. SHAP explanation of XGBoost model classification using typical DPX parameters. **F**. Correlations of the DDM parameters with conventional indices.

A comparison model attempting to classify PwP from controls using conventional measures from the DPX produced a cross-validated AUC of 0.72, indicating worse performance than the DDM-parameter based classification (Figure 4E). The majority of the parameters that had the most utility in differentiating PwP from controls were reaction time measures or indices (AX and BX trial reaction times and PBI-RT). Given its ubiquity in the literature, it was surprising that d’-context was only the 5th most important parameter for differentiating between controls and PwP. Overall, this comparison supports the utility of reaction times and the DDM.

#### Relatives vs. PwP

An XGBoost model classifying PwP from their first-degree relatives produced a cross-validated AUC of 0.82, indicating high discrimination performance. The SHAP analysis (Figure 4B) found that the most important variable in this model was the drift rate on BY trials, v-BY, where slower drift rates predicted a greater likelihood of PwP membership. Other important variables included a-BY, where decreased BY thresholds predicted greater likelihood of being labeled PwP, t-BY, where longer non-decision times for BY trials predicted higher likelihoods of being labeled PwP, t-AY, where longer AY non-decision time predicted higher likelihood of being labeled PwP, and w-A, where decreased A-cue bias predicted higher likelihoods of being labeled PwP. A comparison model attempting to classify PwP from their first-degree relatives using conventional measures from the DPX produced a cross-validated AUC of 0.63, indicating a substantial decrease in performance by comparison to the DDM parameters.

#### Controls vs. Relatives

An XGBoost model classifying controls from relatives produced a cross-validated AUC of 0.82, indicating high discrimination performance. The SHAP analysis (Figure 4C) found that the most important variable in this model was a-BY, where increased BY thresholds predicted higher likelihood of relative membership. Other important variables included w-B, where increased B-cue bias predicted higher likelihoods of being labeled relatives, t-AY, where longer AY non-decision time predicted higher likelihood of being labeled relatives, a-BX, where increased BX thresholds predicted higher likelihood of being labeled relatives, and w-A, where decreased A-cue bias predicted higher likelihoods of being labeled relatives. A comparison model attempting to classify relatives from control using traditional behavioral indices from the DPX produced a cross-validated AUC of 0.65, indicating a substantial decrease in performance by comparison to the DDM parameters.

#### Summary of Comparisons

While the exact ordering of the most useful to least useful parameters found by the SHAP analysis for differentiating between groups varied, there were some commonalities across them (Figure 4D). The most useful parameters for categorizing group membership were the decision threshold and drift rate on BY trials, the non-decision time on AY trials, and the bias on trials with B-cues.

### Relationship of DDM Parameters with Conventional Indices

When examining the correlations between conventional DPX measures and DDM parameters, we found that they were highly associated with each other (Figure 4F). d’-context scores were positively correlated with drift-rate (Kendall’s τ_vAX_=.36, *p*_*adj*_<.001; τ_vAY_=.24, *p*_*adj*_<.001; τ_vBX_=.57, *p*_*adj*_<.001; τ_vBY_=.39, *p*_*adj*_<.001) and degree of bias (τ_zA_=.15, *p*_*adj*_=.03; τ_zB_=.20, *p*_*adj*_=.004). They were also positively correlated with decision threshold in AX and BX trials (τ_aAX_=.15, *p*_*adj*_=.03; τ_aBX_=.17, *p*_*adj*_=.01), and negatively correlated with non-decision time in BX trials (τ_tBX_=-.19, *p*_*adj*_=.005). PBI-RT was significantly correlated with parameters extracted from trials with a B stimulus which assess proactive control (τ_aBX_=-.23, *p*_*adj*_<.001; τ_aBY_=-.31, *p*_*adj*_<.001; τ_vBX_=.44, *p*_*adj*_<.001; τ_vBY_=.26, *p*_*adj*_<.001; τ_tBX_=-.28, *p*_*adj*_<.001; τ_tBY_=-.28, *p*_*adj*_<.001; τ_zB_=-.44, *p*_*adj*_<.001). PBI-RT was also negatively correlated with non-decision time in AY trials (τ_tAY_=-.16, *p*_*adj*_=.02).

## Discussion

We applied a hierarchical drift diffusion model (hDDM) to behavior on the DPX task and found that it provided novel insights into cognitive control deficits in psychosis. The DDM revealed that cognitive control deficits in psychosis are mainly due to two differences in the underlying decision process. First, the slower drift rates across AX, BX, and BY trials suggests a deficit in integrating evidence to respond when the cue stimulus is informative. This is further emphasized by the absence of a similar deficit on AY trials, which can only be responded to correctly and efficiently by refraining from a response based on the cue stimulus. Second, we observed longer non-decision times for PwP during all but AX trials. The slowed non-decision time on proactive control trials (i.e., BX and BY trials) suggests that PwP take longer to represent the probe stimulus or access working memory of the B cue since they had ample time during the interstimulus-interval to anticipate and plan their motor response. The much longer non-decision time on reactive control trials (i.e., AY trials) suggests that PwP have an additional deficit in overriding or generating a new motor response and/or visually recognizing the stimulus when the probe stimulus is unlikely.

While we hypothesized a reduced bias in PwP due to previously observed deficits in proactive control and simulation of the underlying process (Calvin & Redish, 2021), we found mixed observations. PwP showed marginally lower bias than their relatives, even after controlling for age and sex differences, but they were not significantly different from controls. It could be that we failed to detect a difference in degree of bias due to the heterogeneity of our PwP sample. Evidence for this is that the XGBoost classification method found that *increased* bias differentiated PwP from controls, but only for a subset of participants (Figure 4A). The clumps of points in the SHAP analysis suggest that there are subgroups of participants that can be categorized by one or a couple of the parameters. For comparison, the traditional measure classification (Figure 4E) seems to differentiate between the groups in a more continuous fashion. It may be that PwP generally have a deficit in bias as was captured by the LMER but that there is a subset of PwP who engage in a higher degree of preplanning. This suggests that the hDDM is more capable of parsing underlying cognitive processes, and that there may be utility in using it to identify subtypes within psychosis.

Our DDM findings of decreased drift rates and slower non-decision time in PwP are consistent with previous findings on sustained attention, coding, punishment, and reward anticipation tasks (Fish et al., 2018; Mathias et al., 2017; Moustafa et al., 2015; Smucny, Hanks, Lesh, O’Reilly, et al., 2023). Our findings further extend the typically observed inefficiencies in information processing to cognitive control, especially on those trials where responses could be proactively planned. Importantly, our study identified that slowed motor and perceptual speeds appear to be a consistent deficit in people with psychosis. As with previous studies (Fish et al., 2018; Mathias et al., 2017; Smucny, Hanks, Lesh, O’Reilly, et al., 2023), we did not observe differences in the decision thresholds. It may be that the greater decision threshold along with slower drift rates observed by Moustafa et al (2015) is a method of responding more accurately than quickly under sufficient motivation (e.g., during reward or punishment) on a challenging task. Also, the Smucny et al. (2023) analysis approach to AX-CPT behavior was unable to resolve important differences in bias and non-decision time. Based on ours and others findings, their approach of limiting DDM variation to *only* the drift rate and boundary incompletely characterizes the aberrations in cognitive control processes in psychosis.

Relatives of people with psychosis in our study more closely resembled controls than their affected first-degree relatives with psychosis. Their response patterns did not differ statistically from controls on measures of error rates, reaction time, or derived conventional measures. While they also demonstrated comparable drift rates, decision thresholds, and response biases to controls, they did differ from controls in non-decision time on reactive control AY trials. Our patient and relative sample is slightly more heterogeneous in diagnosis than Fish et al.’s sample (2018), but we partially matched their finding of decreased drift rates and increased non-decision times in siblings of participants with schizophrenia compared to controls. Regardless, our XGBoost classification method was able to differentiate relatives from their family members with psychosis, indicating that there are subtle differences in their cognitive control processes from controls. Future studies are needed to better understand the genetic liability of psychotic psychopathology in various decision making processes.

Conventional indices of cognitive control on the DPX were less useful for differentiating between groups than we expected and had interesting relationships with the DDM parameters. d’-context was most correlated with DDM parameters on BX trials, but there are also significant correlations with the drift-rate on trials with other stimulus sequences. Since the drift rate parameter captures error rates more than the other parameters, this suggests that d’-context is measuring general error rates and may be less specific to contextual processing. It is also surprising that d’-context was only the 5th most useful parameter for differentiating between controls and PwP, given its theoretical underpinnings and its ubiquity in the literature. PBI-RT was more informative for differentiating between groups than d’-context and tapped into different aspects of the cognitive control process. Given its basis in reaction times, PBI-RT captured the non-decision time differences that our analysis, and existing literature, suggests are important. Furthermore, the PBI-RT was primarily related to DDM parameters of trials with a B cue. This suggests that it is a very good measure of proactive control, but that it does not differentiate between the underlying mechanisms of the cognitive process. Conventional indices seem to lack an index that specifically taps into the non-decision time on AY trials which had important utility in our group membership classification.

One limitation of this study is that we had to use hierarchical DDMs to estimate individual parameters, which uses information from the group-level to influence parameters at the participant-level. This approach could have caused our XGBoost method to overestimate the area under the curve compared to other grouping approaches. Attempting to fit all participants as a single group is also problematic, however, because it would obscure group differences by assuming a homogeneity across groups that may not exist, and attempting to examining group after fitting each individual’s behavior without group influence is impossible on this task due to the limited number of trials for each subject. To overcome this limitation, it would be beneficial to conduct an experiment with an order of magnitude more trials to estimate the parameters of individual subjects without the influence of other participants. This would permit a better assessment of how informative the DDM parameters are for classification. Within this experiment, it is best to interpret the XGBoost and SHAP results as the order of the most useful parameters for categorization and to be somewhat cautious of the AUC estimates.

Overall, our results provide invaluable information about underlying cognitive control mechanisms that are not captured by conventional measures. By analyzing reaction times from a DDM perspective, we learned that slowed motor/perceptual time as well as inefficiencies in proactive information integration likely contribute to deficits in cognitive control in psychosis. Our findings provide additional support for a deficit in proactive control in psychotic psychopathology, and further highlight the importance of perceptual and motor functions in understanding compromised cognitive control in people with a history of psychosis.

## Methods

### Participants and Clinical Measures

Two-hundred-and-fifty-three participants were recruited as part of the Psychosis Human Connectome Project. For a detailed description of the Psychosis Human Connectome Project, please see Demro et al., (2021). In summary, 123 people with psychosis (PwP), 79 of their first-degree biological relatives, and 51 controls were included in the analysis. All participants completed a clinical interview and self-report questionnaires, as well as cognitive and motor assessments. Trained research assistants conducted the Structured Clinical Interview for DSM-IV-TR disorders (First et al., 2002), and the Psychosis Module of the Diagnostic Interview for Genetic Studies (DIGS; Nurnberger, 1994) with each of the participants to obtain diagnostic information. Diagnostic consensus was completed by a team of at least two qualified assessors (clinical psychology graduate students, postdoctoral associates, or licensed psychologists) to determine which diagnostic criteria were met, and reached consensus on the most appropriate DSM diagnoses. Among the 123 PwP, 73 were individuals with schizophrenia or schizophreniform, 15 were individuals with schizoaffective disorder, and 35 were individuals with bipolar disorder with psychosis. In addition to making diagnostic determinations, we also collected symptomatology measures. The Brief Psychiatric Rating Scale-24 Item Version (Lukoff et al., 1986; Wilson & Sponheim, 2014) and the Scales for the Assessment of Negative/Positive Symptoms (Andreasen, 1981, 1983) were used to assess the participants’ psychotic, depressive, and manic symptoms for the 30 days leading up to the evaluation based on the participants recollection. Table 1 presents the demographics information of the participants.

### The Dot Pattern Expectancy (DPX) Task and Conventional Measures

We used the dot-pattern version of the AX-CPT cognitive control task, the DPX task, where stimuli were braille-based arrangements of dots (MacDonald et al., 2005). On each trial, a cue stimulus and a probe stimulus were sequentially presented with an interstimulus interval of 2500 or 3500 ms between them (Figure 1A). Cue and probe stimuli were differentiable by being colored white and light blue, respectively. Stimuli were grouped into ‘valid’ and ‘invalid’ categories and only when both the cue and probe were valid was the participant supposed to provide a ‘target’ response with the index finger of their right hand. The only target sequence was the A cue followed by the X probe, and all other cue-probe combinations were considered non-target (Figure 1A). As such, there were four cue-probe permutations: AX, AY, BX, and BY. The participant was to provide a non-target response with the middle finger of the same hand for any non-target sequence (i.e., AY, BX, or BY). An expectation bias was induced by having 60% of the cue-probe trials being the target sequence (AX). The remaining trials were distributed such that 15% of trials were AY, 15% BX, and 10% BY cue-probe pairings. Each participant responded to 120 trials that were equally distributed across 3 blocks. While not analyzed in this manuscript, the participants were undergoing fMRI during this task. About 2% of all trials were responded to within 100 ms (too fast to have recognized the probe stimulus) or were not responded to within 2000 ms, and were thus excluded from subsequent analyses.

### d’-context and Proactive Behavioral Index (BPI) Calculation

A number of conventional measures have been used in the DPX literature to assess cognitive control. d’-context scores, a measure of context processing, were calculated as the difference between correct responses to the AX pair and incorrect responses to the BX pair: z(AX_Hit_)-z(BX_FalseAlarm_) (Servan-Schreiber et al., 1996). Higher d’-context scores are suggestive of better context processing. PBI in DPX for accuracy and reaction time (Braver et al., 2009), a measure of proactiveness, was calculated by (AY-BX)/(AY+BX). This measure evaluates proactiveness by directly comparing error rates or reaction time of proactive vs. reactive trial types. Higher PBIs suggest a preference for proactive control over reactive control. Due to poor psychometric properties (excessively kurtotic with large numbers of outliers), we did not relate the PBI-Accuracy with the hDDM parameters.

### Hierarchical Drift Diffusion Model (hDDM)

Hierarchical DDMs were applied to the DPX task data across groups using the `HDDM` 0.9.1 python package (Wiecki et al., 2013). Since we had *a priori* reasons to believe that the evidence integration process would vary across diagnostic categories, we fitted the hDDMs to each group separately. To determine the best description of the participant behavior, we fitted several models that varied whether the parameter values of *a, t*, and *v* parameters were consistent across trial sequences. We decided to only permit *w* to vary by the cue stimulus because it theoretically should only influence the start of the evidence integration process and should, thus, not be modified by the probe stimulus. We identified the best-fit models by comparing the deviance information criteria (DIC) and by evaluating how well the model converged. Model convergence was assessed via the Gelman-Rubin ^*R*statistic (Gelman et al., 2013).

### Conventional and DDM Statistical Analyses

All statistical analyses were conducted in `R` (version 4.2.3; R Core Team, 2023) and corrected for age, biological sex, and family-level dependencies. Conventional variables from DPX, including error rates and correct trial reaction time, and DDM parameters were examined by Linear Mixed-Effect Regression models (LMER) using the `lme4` package (version 1.1-31; Bates et al., 2015). Post-hoc analyses examining interaction and group effects were performed via the `emmeans` package (version 1.8.3; Lenth, 2022) with multivariate *t*-distribution adjustments for multiple comparisons. d’-context scores and PBIs were analyzed with one-way ANOVAs, including post-hoc group comparisons corrected by multivariate *t*-distributions. Since most of the conventional measures were non-Gaussian, correlational analyses between conventional variables and DDM parameters were performed with the robust, nonparametric Kendall’s Rank Correlation Tests (Kendall, 1938). We controlled for the false discovery rate when making multiple family-wise correlational tests (Benjamini & Hochberg, 1995).

### eXtreme Gradient Boosting (XGBoost) Classifier and Shapley Value Model Explanation

To determine the multivariate predictive utility of DDM parameters, we used a nonparametric machine learning classification approach called eXtreme Gradient Boosting, or XGBoost (Chen & Guestrin, 2016), implemented in the R package `xgboost` version 1.7.5. This classifier is more stable with a smaller sample size than competing methods like support vector machines (Floares et al., 2017; Mukherjee et al., 2003). Additionally, this classifier is robust to class imbalance (Wang et al., 2020), which is ideal because our groups varied in sizes. We fit three classifiers to classify PwP from controls, PwP from first-degree relatives, and first-degree relatives from controls. We fit another three classifiers to classify across the same groups but with conventional behavioral indices from the DPX: accuracy and RT of all four trial types and the PBI-RT, PBI-accuracy, and d’-context. All XGBoost analyses used the area under the receiver operating characteristic curve (AUC) as the objective function. The AUC can range from 0 to 1, where 1 is perfect classification and 0.5 indicates completely random performance for a binary outcome.

XGBoost requires several hyperparameters. For brevity, we do not describe the interpretation of each hyperparameter. It is recommended to tune hyperparameters in a data-driven way (Bentéjac et al., 2021). We used a Bayesian hyperparameter tuning approach with 100 repetitions implemented in the `mlrMBO` package for R version 1.1.5.1 (Bischl et al., 2018) to tune XGBoost hyperparameters. For each classifier, we tuned parameters in 25% of the full sample who were not further analyzed, ensuring that hyperparameter optimization occurred in a different group of subjects than our primary analysis. This reduces risk of model overfitting. Hyperparameters were optimized over the Cartesian product of: number of rounds = [50, 100, 150, 200], max tree depth = [3, 4, 5, 6, 7, 8, 9, 10], eta = [0.01, 0.03, 0.1, 0.3], gamma = [0, 0.1, 0.5, 1, 5], subsampling = [0.5, 0.6, 0.7, 0.8, 0.9, 1], minimum child weight = [1, 3, 5].

Following hyperparameter tuning in 25% of the sample, we used XGBoost with optimized hyperparameters to classify participants in the remaining 75% of the sample. We trained and tested classifiers using 5-fold cross-validation, ensuring that all machine learning models were trained and tested on fully independent sets of data, which also reduced the risk of model overfitting. During cross-validation of the model that classified PwP from their first-degree relatives, we always assigned family members to the same cross-validation fold, ensuring that models were not overfit by training and testing on related family members. For this analysis, we report the final cross-validated AUC for each classifier as our primary outcome.

XGBoost provides a powerful tool for machine learning classification, but multivariate machine learning models like XGBoost can suffer from reduced interpretability due to the black-box nature of these models and the complicated model fitting process. We solved this problem by applying a recently developed model explanation tool to the output of each XGBoost classifier, using the R package `SHAPforxgboost` version 0.1.1 (Just et al., 2020). SHapley Additive exPlanations, or SHAP (Lundberg et al., 2019; Lundberg & Lee, 2017), is an information theoretic approach that explains the output of a machine learning model by ranking the input variables according to which variables had the greatest independent contribution to the output of the classifier. SHAP is based on the game theoretic concept of Shapley values, proposed by (Shapley, 1953) as a consistent way to allocate credit to a team of players working toward a common goal. In this case, each independent variable is treated as a player on the team, and the common goal is to maximize the performance of the classifier. As such, this analysis provides an interpretation for the output of the XGBoost algorithm, and describes which DDM parameters are most important for discriminating between participant groups.

## Data Availability

All data produced in the present study in the present study are available upon reasonable request to the authors.

## Acknowledgements

We would like to thank Angus MacDonald III who provided helpful feedback on an earlier version of this manuscript. This research was supported by the following grants: National Institute of Drug Abuse T32DA037183 (O.L. Calvin), National Institutes of Health’s National Center for Advancing Translational Sciences TL1TR002493 (E. Rawls) and UL1TR002494 (E. Rawls), and National Institutes of Health grants U01MH108150 (S.R. Sponheim) and P50MH119569 (A.D. Redish).

## REFERENCES

Alptekin, K., Akvardar, Y., Akdede, B. B. K., Dumlu, K., Işık, D., Pirinçci, F., Yahssin, S., & Kitiş, A. (2005). Is quality of life associated with cognitive impairment in schizophrenia? Progress in Neuro-Psychopharmacology and Biological Psychiatry, 29(2), 239–244. https://doi.org/10.1016/j.pnpbp.2004.11.006

Andreasen, N. C. (1981). The Scale for the Assessment of Negative Symptoms (SANS).

Andreasen, N. C. (1983). The Scale for the Assessment of Positive Symptoms (SAPS).

Bates, D., Mächler, M., Bolker, B., & Walker, S. (2015). Fitting Linear Mixed-Effects Models Using lme4. Journal of Statistical Software, 67(1). https://doi.org/10.18637/jss.v067.i01

Benjamini, Y., & Hochberg, Y. (1995). Controlling the False Discovery Rate: A Practical and Powerful Approach to Multiple Testing. Journal of the Royal Statistical Society: Series B (Methodological), 57(1), 289–300. https://doi.org/10.1111/j.2517-6161.1995.tb02031.x

Bentéjac, C., Csörgő, A., & Martínez-Muñoz, G. (2021). A Comparative Analysis of XGBoost. Artificial Intelligence Review, 54(3), 1937–1967. https://doi.org/10.1007/s10462-020-09896-5

Bischl, B., Richter, J., Bossek, J., Horn, D., Thomas, J., & Lang, M. (2018). mlrMBO: A Modular Framework for Model-Based Optimization of Expensive Black-Box Functions (arXiv:1703.03373). arXiv. http://arxiv.org/abs/1703.03373

Braver, T. S. (2012). The variable nature of cognitive control: A dual mechanisms framework. Trends in Cognitive Sciences, 16(2), 106–113. https://doi.org/10.1016/j.tics.2011.12.010

Braver, T. S., Barch, D. M., Keys, B. A., Carter, C. S., Cohen, J. D., Kaye, J. A., Janowsky, J. S., Taylor, S. F., Yesavage, J. A., Mumenthaler, M. S., Jagust, W. J., & Reed, B. R. (2001). Context processing in older adults: Evidence for a theory relating cognitive control to neurobiology in healthy aging. Journal of Experimental Psychology: General, 130(4), 746–763. https://doi.org/10.1037/0096-3445.130.4.746

Braver, T. S., Paxton, J. L., Locke, H. S., & Barch, D. M. (2009). Flexible neural mechanisms of cognitive control within human prefrontal cortex. Proceedings of the National Academy of Sciences, 106(18), 7351–7356. https://doi.org/10.1073/pnas.0808187106

Calvin, O. L., & Redish, A. D. (2021). Global disruption in excitation-inhibition balance can cause localized network dysfunction and Schizophrenia-like context-integration deficits. PLOS Computational Biology, 17(5), e1008985. https://doi.org/10.1371/journal.pcbi.1008985

Chen, T., & Guestrin, C. (2016). XGBoost: A Scalable Tree Boosting System. Proceedings of the 22nd ACM SIGKDD International Conference on Knowledge Discovery and Data Mining, 785–794. https://doi.org/10.1145/2939672.2939785

Cohen, J. D., Barch, D. M., Carter, C., & Servan-Schreiber, D. (1999). Context-processing deficits in schizophrenia: Converging evidence from three theoretically motivated cognitive tasks. Journal of Abnormal Psychology, 108(1), 120–133. https://doi.org/10.1037/0021-843X.108.1.120

Delawalla, Z., Csernansky, J. G., & Barch, D. M. (2008). Prefrontal Cortex Function in Nonpsychotic Siblings of Individuals with Schizophrenia. Biological Psychiatry, 63(5), 490–497. https://doi.org/10.1016/j.biopsych.2007.05.007

Demro, C., Mueller, B. A., Kent, J. S., Burton, P. C., Olman, C. A., Schallmo, M.-P., Lim, K. O., & Sponheim, S. R. (2021). The psychosis human connectome project: An overview. NeuroImage, 241, 118439. https://doi.org/10.1016/j.neuroimage.2021.118439

Eriksen, B. A., & Eriksen, C. W. (1974). Effects of noise letters upon the identification of a target letter in a nonsearch task. Perception & Psychophysics, 16(1), 143–149. https://doi.org/10.3758/BF03203267

First, M. B., Spitzer, R. L., Gibbon, M., & Williams, J. B. (2002). Structured Clinical Interview for DSM-IV-TR Axis I Disorders, Research Version, Patient Edition.

Fish, S., Toumaian, M., Pappa, E., Davies, T. J., Tanti, R., Saville, C. W. N., Theleritis, C., Economou, M., Klein, C., & Smyrnis, N. (2018). Modelling reaction time distribution of fast decision tasks in schizophrenia: Evidence for novel candidate endophenotypes. Psychiatry Research, 269, 212–220. https://doi.org/10.1016/j.psychres.2018.08.067

Floares, A. G., Ferisgan, M., Onita, D., Ciuparu, A., Calin, G. A., & Manolache, F. B. (2017). The Smallest Sample Size for the Desired Diagnosis Accuracy. International Journal of Oncology and Cancer Therapy, 2, 13–19.

Gelman, A., Carlin, J. B., Stern, H. S., Dunson, D. B., Vehtari, A., & Rubin, D. B. (2013). Bayesian Data Analysis (0 ed.). Chapman and Hall/CRC. https://doi.org/10.1201/b16018

Gupta, A., Bansal, R., Alashwal, H., Kacar, A. S., Balci, F., & Moustafa, A. A. (2022). Neural Substrates of the Drift-Diffusion Model in Brain Disorders. Frontiers in Computational Neuroscience, 15, 678232. https://doi.org/10.3389/fncom.2021.678232

Jones, J. A. H., Sponheim, S. R., & MacDonald, A. W. (2010). The dot pattern expectancy task: Reliability and replication of deficits in schizophrenia. Psychological Assessment, 22(1), 131–141. https://doi.org/10.1037/a0017828

Just, A. C., Liu, Y., Sorek-Hamer, M., Rush, J., Dorman, M., Chatfield, R., Wang, Y., Lyapustin, A., & Kloog, I. (2020). Gradient boosting machine learning to improve satellite-derived column water vapor measurement error. Atmospheric Measurement Techniques, 13(9), 4669–4681. https://doi.org/10.5194/amt-13-4669-2020

Kendall, M. G. (1938). A New Measure of Rank Correlation. Biometrika, 30, 81–93.

Lam, N. H., Borduqui, T., Hallak, J., Roque, A., Anticevic, A., Krystal, J. H., Wang, X.-J., & Murray, J. D. (2022). Effects of Altered Excitation-Inhibition Balance on Decision Making in a Cortical Circuit Model. The Journal of Neuroscience, 42(6), 1035–1053. https://doi.org/10.1523/JNEUROSCI.1371-20.2021

Lenth, R. (2022). emmeans: Estimated Marginal Means, aka Least-Squares Means (1.8.3) [Computer software]. https://CRAN.R-project.org/package=emmeans

Lesh, T. A., Westphal, A. J., Niendam, T. A., Yoon, J. H., Minzenberg, M. J., Ragland, J. D., Solomon, M., & Carter, C. S. (2013). Proactive and reactive cognitive control and dorsolateral prefrontal cortex dysfunction in first episode schizophrenia. NeuroImage: Clinical, 2, 590–599. https://doi.org/10.1016/j.nicl.2013.04.010

Limongi, R., Bohaterewicz, B., Nowicka, M., Plewka, A., & Friston, K. J. (2018). Knowing when to stop: Aberrant precision and evidence accumulation in schizophrenia. Schizophrenia Research, 197, 386–391. https://doi.org/10.1016/j.schres.2017.12.018

Lopez-Garcia, P., Young Espinoza, L., Molero Santos, P., Marin, J., & Ortuño Sanchez-Pedreño, F. (2013). Impact of COMT genotype on cognition in schizophrenia spectrum patients and their relatives. Psychiatry Research, 208(2), 118–124. https://doi.org/10.1016/j.psychres.2012.09.043

Lukoff, D., Liberman, R. P., & Nuechterlein, K. H. (1986). Symptom Monitoring in the Rehabilitation of Schizophrenic Patients. Schizophrenia Bulletin, 12(4), 578–603. https://doi.org/10.1093/schbul/12.4.578

Lundberg, S. M., Erion, G. G., & Lee, S.-I. (2019). Consistent Individualized Feature Attribution for Tree Ensembles. ArXiv.

Lundberg, S. M., & Lee, S.-I. (2017). A Unified Approach to Interpreting Model Predictions. Advances in Neural Information Processing Systems, 4765–4774.

MacDonald, A. W., & Carter, C. S. (2003). Event-Related fMRI Study of Context Processing in Dorsolateral Prefrontal Cortex of Patients With Schizophrenia. Journal of Abnormal Psychology, 112(4), 689–697. https://doi.org/10.1037/0021-843X.112.4.689

MacDonald, A. W., Goghari, V. M., Hicks, B. M., Carter, C. S., Flory, J. D., & Manuck, S. B. (2005). A Convergent–Divergent Approach to Context Processing, General Intellectual Functioning, and the Genetic Liability to Schizophrenia. Neuropsychology, 19(6), 8.

MacDonald, A. W., Pogue-Geile, M. F., Johnson, M. K., & Carter, C. S. (2003). A Specific Deficit in Context Processing in the Unaffected Siblings of Patients With Schizophrenia. Archives of General Psychiatry, 60(1), 57. https://doi.org/10.1001/archpsyc.60.1.57

Mathias, S. R., Knowles, E. E. M., Barrett, J., Leach, O., Buccheri, S., Beetham, T., Blangero, J., Poldrack, R. A., & Glahn, David. C. (2017). The Processing-Speed Impairment in Psychosis Is More Than Just Accelerated Aging. Schizophrenia Bulletin, sbw168. https://doi.org/10.1093/schbul/sbw168

Moustafa, A. A., Kéri, S., Somlai, Z., Balsdon, T., Frydecka, D., Misiak, B., & White, C. (2015). Drift diffusion model of reward and punishment learning in schizophrenia: Modeling and experimental data. Behavioural Brain Research, 291, 147–154. https://doi.org/10.1016/j.bbr.2015.05.024

Mukherjee, S., Tamayo, P., Rogers, S., Rifkin, R., Engle, A., Campbell, C., Golub, T. R., & Mesirov, J. P. (2003). Estimating Dataset Size Requirements for Classifying DNA Microarray Data. Journal of Computational Biology, 10(2), 119–142. https://doi.org/10.1089/106652703321825928

Nurnberger, J. I. (1994). Diagnostic Interview for Genetic Studies: Rationale, Unique Features, and Training. Archives of General Psychiatry, 51(11), 849. https://doi.org/10.1001/archpsyc.1994.03950110009002

Pascal de Raykeer, R., Hoertel, N., Blanco, C., Lavaud, P., Kaladjian, A., Blumenstock, Y., Cuervo-Lombard, C.-V., Peyre, H., Lemogne, C., Limosin, F., Adès, J., Alezrah, C., Amado, I., Amar, G., Andréi, O., Arbault, D., Archambault, G., Aurifeuille, G., Barrière, S., … Zendjidjian, X. (2019). Effects of depression and cognitive impairment on quality of life in older adults with schizophrenia spectrum disorder: Results from a multicenter study. Journal of Affective Disorders, 256, 164–175. https://doi.org/10.1016/j.jad.2019.05.063

Poppe, A. B., Barch, D. M., Carter, C. S., Gold, J. M., Ragland, J. D., Silverstein, S. M., & MacDonald, A. W. (2016). Reduced Frontoparietal Activity in Schizophrenia Is Linked to a Specific Deficit in Goal Maintenance: A Multisite Functional Imaging Study. Schizophrenia Bulletin, 42(5), 1149–1157. https://doi.org/10.1093/schbul/sbw036

Poppe, A. B., Carter, C. S., Minzenberg, M. J., & MacDonald, A. W. (2015). Task-based functional connectivity as an indicator of genetic liability to schizophrenia. Schizophrenia Research, 162(1–3), 118–123. https://doi.org/10.1016/j.schres.2014.11.022

R Core Team. (2023). R: A language and environment for statistical computing. [Computer software]. R Foundation for Statistical Computing. https://www.R-project.org/.

Reilly, J. L., Hill, S. K., Gold, J. M., Keefe, R. S. E., Clementz, B. A., Gershon, E., Keshavan, M. S., Pearlson, G., Tamminga, C. A., & Sweeney, J. A. (2016). Impaired Context Processing is Attributable to Global Neuropsychological Impairment in Schizophrenia and Psychotic Bipolar Disorder. Schizophrenia Bulletin. https://doi.org/10.1093/schbul/sbw081

Richard, A. E., Carter, C. S., Cohen, J. D., & Cho, R. Y. (2013). Persistence, diagnostic specificity and genetic liability for context-processing deficits in schizophrenia. Schizophrenia Research, 147(1), 75–80. https://doi.org/10.1016/j.schres.2013.02.020

Savilla, K., Kettler, L., & Galletly, C. (2008). Relationships Between Cognitive Deficits, Symptoms and Quality of Life in Schizophrenia. Australian & New Zealand Journal of Psychiatry, 42(6), 496–504. https://doi.org/10.1080/00048670802050512

Servan-Schreiber, D. (1996). Schizophrenic Deficits in the Processing of Context: A Test of a Theoretical Model. Archives of General Psychiatry, 53(12), 1105. https://doi.org/10.1001/archpsyc.1996.01830120037008

Shapley, L. S. (1953). A Value for N-Person Games. In Contributions to the Theory of Games (2nd ed., Vol. 28, pp. 307–317). Princeton University Press.

Smucny, J., Barch, D. M., Gold, J. M., Strauss, M. E., MacDonald, A. W., Boudewyn, M. A., Ragland, J. D., Silverstein, S. M., & Carter, C. S. (2019). Cross-diagnostic analysis of cognitive control in mental illness: Insights from the CNTRACS consortium. Schizophrenia Research, 208, 377–383. https://doi.org/10.1016/j.schres.2019.01.018

Smucny, J., Hanks, T. D., Lesh, T. A., & Carter, C. S. (2023). Altered Associations between Task Performance and Dorsolateral Prefrontal Cortex Activation during Cognitive Control in Schizophrenia. Biological Psychiatry: Cognitive Neuroscience and Neuroimaging, S2451902223001301. https://doi.org/10.1016/j.bpsc.2023.05.010

Smucny, J., Hanks, T. D., Lesh, T. A., O’Reilly, R. C., & Carter, C. S. (2023). Altered Associations Between Motivated Performance and Frontostriatal Functional Connectivity During Reward Anticipation in Schizophrenia. Schizophrenia Bulletin, 49(3), 717–725. https://doi.org/10.1093/schbul/sbac204

Stephenson, D. D., El Shaikh, A. A., Shaff, N. A., Bustillo, J. R., Dodd, A. B., Wertz, C. J., Ryman, S. G., Hanlon, F. M., Hogeveen, J. P., Ling, J. M., Yeo, R. A., Stromberg, S. F., Lin, D. S., Abrams, S., & Mayer, A. R. (2020). Differing functional mechanisms underlie cognitive control deficits in psychotic spectrum disorders. Journal of Psychiatry and Neuroscience, 45(6), 430–440. https://doi.org/10.1503/jpn.190212

Stroop, J. R. (1935). Studies of Interference in Serial Verbal Reactions. Journal of Experimental Psychology, 18, 643–662.

Ueoka, Y., Tomotake, M., Tanaka, T., Kaneda, Y., Taniguchi, K., Nakataki, M., Numata, S., Tayoshi, S., Yamauchi, K., Sumitani, S., Ohmori, T., Ueno, S., & Ohmori, T. (2011). Quality of life and cognitive dysfunction in people with schizophrenia. Progress in Neuro-Psychopharmacology and Biological Psychiatry, 35(1), 53–59. https://doi.org/10.1016/j.pnpbp.2010.08.018

Wang, C., Deng, C., & Wang, S. (2020). Imbalance-XGBoost: Leveraging weighted and focal losses for binary label-imbalanced classification with XGBoost. Pattern Recognition Letters, 136, 190–197. https://doi.org/10.1016/j.patrec.2020.05.035

Wiecki, T. V., Sofer, I., & Frank, M. J. (2013). HDDM: Hierarchical Bayesian estimation of the Drift-Diffusion Model in Python. Frontiers in Neuroinformatics, 7. https://doi.org/10.3389/fninf.2013.00014

Wilson, S., & Sponheim, S. R. (2014). Dimensions underlying psychotic and manic symptomatology: Extending normal-range personality traits to schizophrenia and bipolar spectra. Comprehensive Psychiatry, 55(8), 1809–1819. https://doi.org/10.1016/j.comppsych.2014.07.008

